# Environmental equity and COVID-19 experiences in the United States: Results from three survey waves of a nationally representative study conducted between 2020-2022

**DOI:** 10.1101/2023.05.16.23290050

**Authors:** Elyssa Anneser, Thomas J. Stopka, Elena N. Naumova, Keith R. Spangler, Kevin J. Lane, Andrea Acevedo, Jeffrey K. Griffiths, Yan Lin, Peter Levine, Laura Corlin

**Author notes:** Corresponding author: Laura Corlin, 136 Harrison Avenue, Boston, MA 02111. Data sharing statement: Analytic code is available upon reasonable request from the corresponding author. Individual-level survey data may be made available upon reasonable request by email to Thomas Stopka.

## Abstract

Certain environmental exposures, such as air pollution, are associated with COVID-19 incidence and mortality. To determine whether environmental context is associated with other COVID-19 experiences, we used data from the nationally representative Tufts Equity in Health, Wealth, and Civic Engagement Study data (n=1785; three survey waves 2020-2022). Environmental context was assessed using self-reported climate stress and county-level air pollution, greenness, toxic release inventory site, and heatwave data. Self-reported COVID-19 experiences included willingness to vaccinate against COVID-19, health impacts from COVID-19, receiving assistance for COVID-19, and provisioning assistance for COVID-19. Self-reported climate stress in 2020 or 2021 was associated with increased COVID-19 vaccination willingness by 2022 (odds ratio [OR] = 2.35; 95% confidence interval [CI] = 1.47, 3.76), even after adjusting for political affiliation (OR = 1.79; 95% CI = 1.09, 2.93). Self-reported climate stress in 2020 was also associated with increased likelihood of receiving COVID-19 assistance by 2021 (OR = 1.89; 95% CI = 1.29, 2.78). County-level exposures (i.e., less greenness, more toxic release inventory sites, more heatwaves) were associated with increased vaccination willingness. Air pollution exposure in 2020 was positively associated with likelihood of provisioning COVID-19 assistance in 2020 (OR = 1.16 per µg/m^3^; 95% CI = 1.02, 1.32). Associations between certain environmental exposures and certain COVID-19 outcomes were stronger among those who identify as a race/ethnicity other than non-Hispanic White and among those who reported experiencing discrimination; however, these trends were not consistent. A latent variable representing a summary construct for environmental context was associated with COVID-19 vaccination willingness. Our results add to the growing body of literature suggesting that intersectional equity issues affecting likelihood of exposure to adverse environmental conditions are also associated with health-related outcomes.

## Introduction

Over two million COVID-19 cases were diagnosed in the United States (US) within the first five months of the pandemic; nearly 2.5 years into the pandemic, over 80 million cases occurred in the US.^1^ Although the devastation of this pandemic has been felt by everyone, the health, economic, and social consequences have been disproportionately borne by historically minoritized and marginalized communities (e.g., Black/African American, Hispanic/Latinx, and Indigenous communities).^2–13^ Moreover, these same communities have dealt with race-based residential segregation, redlining, and other forms of environmental racism resulting in disproportionately high exposure to worse air quality, less greenspace, more extreme heat events, more toxic chemicals, and more climate stress.^14–21^ Environmental inequities intersect with and exacerbate other forms of structural racism to worsen health disparities. For example, poor air quality, low greenspace exposure, and high Toxics Release Inventory (TRI) site (facilities that release specific pollutants that are harmful to human health and that are required to report the quantity of releases for each chemical annually to the US Environmental Protection Agency) exposure are each associated with worse COVID-19 health outcomes.^22–27^

Despite extensive prior assessment of associations between specific environmental exposures (e.g., air pollution) and population-level COVID-19 incidence and mortality trends,^23, 28^ less is understood about how environmental context affects other COVID-19 experiences. Using individual-level health data to characterize the association between environmental context (accounting for multiple adverse environmental exposures simultaneously) and a larger set of COVID-19 experiences could provide greater insight into the intersection of environmental and other structural inequities. It could inform more equitable and efficient COVID-19 policies and advance pandemic preparedness efforts moving forward.

To address this critical long-term goal, we used three waves of data from a nationally representative survey to investigate associations between environmental exposures and a broad set of COVID-19 experiences (i.e., willingness to vaccinate against COVID-19, COVID-19 health impact, COVID-19 assistance recipient, COVID-19 assistance provider). Our primary aim was to assess associations between individual environmental exposures (county-level annual average air quality, greenness, heat wave days, and TRI sites; self-reported climate stress) and COVID-19 experiences. We had two secondary aims. One was to assess whether the associations observed in the primary aim were modified by race/ethnicity or discrimination experiences. The other was to conduct a preliminary assessment of associations between a summary construct of environmental context and each COVID-19 experience, recognizing that the environmental exposures may not directly lead to the COVID-19 experiences but that exposure to multiple adverse environmental exposures together may serve as a proxy for the experience of environmental inequity.

## Methods

### Study population and survey administration

We used data from the Tufts Equity in Health, Wealth, and Civic Engagement Study. This study was approved by the Tufts Institutional Review Board (protocol STUDY00000428). The study uses a survey that has been conducted over three waves. The survey methods for the first two waves have been described previously.^29^ Briefly, the study had a target population of all non-institutionalized adults residing in the US. To reach a nationally representative sample of this population, Ipsos Public Affairs administered a survey to members of its KnowledgePanel. This panel was developed in 1999 and included participants who were recruited using probability-based sampling techniques. To ensure continued representativeness of the geodemographic composition of the adult population of the US, Ipsos uses stratified random sampling, provides internet access to invited individuals who otherwise do not have access, and includes Spanish versions of the surveys. Once invited individuals are recruited and complete a Core Profile Survey, they become eligible for selection into subsequent surveys, such as the Tufts Equity in Health, Wealth, and Civic Engagement Study. Participants earn standard incentive payment from Ipsos for completing surveys (the cash equivalent of approximately $1 for our survey and entrance into larger cash prize sweepstakes).

The Tufts Equity in Health, Wealth, and Civic Engagement Study included data from 2545 participants collected in up to three survey waves (May 29-June 10, 2020; April 23-May 3, 2021; May 26-June 2, 2022). In the initial wave, a random sample of 1980 Ipsos KnowledgePanel members were invited to participate. Of these individuals, 1267 individuals responded for a 64% final stage completion rate. Over 99% of the participants in the first wave of the survey answered the demographic surveys in March 2019 or later, and >95% answered all health-related questions in January 2019 or later. All individuals who completed the first wave of the survey were invited to complete the second wave of the survey, and 931 did (73% retention). Additionally, 840 individuals who identify as non-Hispanic Black/African American or as Hispanic of any race were invited as part of an over-sample for wave 2, and 518 of these individuals participated (62%). In total, there were 1449 participants in wave 2 for a completion rate of 69%. In wave 3, all individuals who completed waves 1 and/or 2 were invited to participate and 1071 did (42% retention: n = 709 with data from all three waves). Additionally, 1304 other individuals who identify as non-Hispanic Black/African American, Hispanic, or non-Hispanic Asian (including Hawaiian/other Pacific Islander) were invited as part of an over-sample and 760 participated (none of whom were included in this analysis since they were not part of the earlier waves of data collection and were therefore missing exposure data). In total, there were 1831 participants who completed wave 3 (66% completion rate). A total of 1785 participants participated in at least one of wave 1 or wave 2, and therefore contributed data for the analyses in this paper (Figure 1).

**Figure 1.**
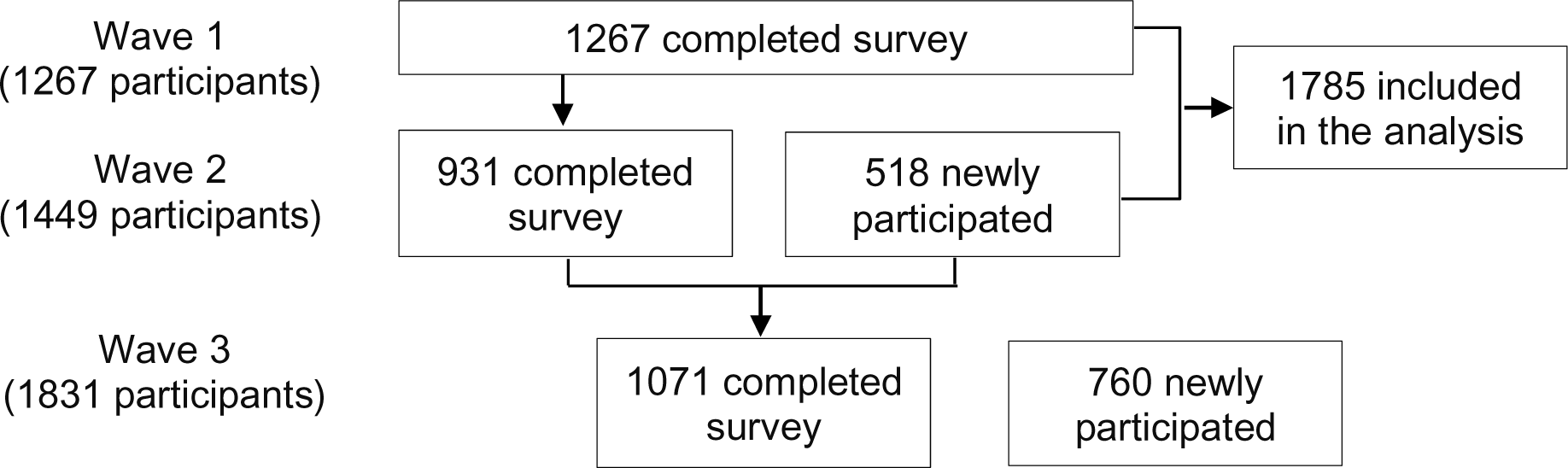
Flow diagram for participation in the study

For each survey wave, study-specific post-stratification weights were calculated based on the Current Population Survey, the US Census Bureau’s American Community Survey, and the weighted KnowledgePanel profile data.^30, 31^ Waves two and three account for design weights for the added participants in the over-sample. All weights accounted for sociodemographic factors such as gender, age group, Census region, metropolitan status, educational attainment, and household income. Survey weights varied from 0.072 to 8.509, with medians of 0.826 at wave 2 and 0.693 at wave 3. The overall design effects for waves 2 and 3 were 1.575 and 2.176, respectively.

### Survey-derived variables

Demographics: Participants self-reported their gender (female/male; although these terms should refer to biologic sex, they were labeled as gender in the survey), age (full years, continuous), educational attainment (less than high school/high school/some college/Bachelor’s or higher), race/ethnicity (Hispanic [any race]/non-Hispanic Black or African American/non-Hispanic White/at least two races or other [including American Indian, Alaska Native, Asian, Native Hawaiian, and Pacific Islander]), and annual household income (<$25,000/$25,000-$49,999/$50,000-$74,999/$75,000-$99,999/$100,000-$149,999/≥$150,000). Political party affiliation was also derived from wave 1 data where available (Democrat/Republican/Independent) or wave 2 data if wave 1 data were unavailable (Democrat [strong Democrat or not strong Democrat]/Republican [strong Republican or not strong Republican]/Independent [leans Democrat or leans Republican or undecided or independent or other]). Residential locations were determined to be in a metropolitan statistical area or not by matching residential addresses to Census Block Groups (or ZIP codes if Census Block Groups were unavailable) and then using the metropolitan statistical area designation for the Census Block Group.

#### Discrimination

In wave 1, participants also indicated whether the following situations had ever occurred to them: “*You have been unfairly stopped, searched, questioned, physically threatened or abused by the police*,” “*Someone you know has been unfairly stopped, searched, questioned, physically threatened or abused by the police*,” “*You were mistaken for someone else of your same race/ethnicity (who may not look like you at all)*,” and “*You have been unfairly prevented from having access to a service or been treated unfairly by a service provider*.” Participants who provided affirmative responses to any of these questions, along with participants who responded that any of the following situations occurred “sometimes” or “frequently” were considered to have experienced discrimination: “*Being treated with less courtesy or respect than other people*,” “*Feeling that people act as if they are afraid of you*,” and “*Receiving poorer service than others in restaurants and stores*.” In wave 2, participants were considered to have experienced discrimination if any of the following situations occurred at least a few times a year: “*You are treated with less courtesy or respect than other people*,” “*You receive poorer service than other people at restaurants or stores*,” “*People act as if they think you are not smart*,” “*People act as if they are afraid of you*,” “*You are threatened or harassed*,” “*You have been unfairly stopped, searched, questioned, physically threatened or abused by the police*,” and “*Someone you know has been unfairly stopped, searched, questioned, physically threatened or abused by the police*.”

#### Exposure and outcomes

One environmental exposure variable (climate stress) and all four COVID-19 outcome variables were based on the survey responses. In wave 1 participants were considered to have climate stress if they responded “somewhat stressful” or “very stressful” to the question, “*How stressful is climate change/global warming for you?*” In wave 2, participants were considered to have climate stress if they moderately or strongly agreed with the statement, “*Thinking about climate change makes me feel anxious*.” The four COVID-19 outcomes for survey waves one and two were willingness to vaccinate against COVID-19, COVID-19 health impact, COVID-19 assistance recipient, and COVID-19 assistance provider. Two of these outcomes were also assessed in wave 3 (willingness to vaccinate against COVID-19 and COVID-19 health impact). We dichotomized each of these variables based on the criteria described in Table 1. When included as a response option, “Don’t know” was grouped with “No” responses. For the COVID-19 health impact variable in wave 1, we included COVID-19 testing given the limited availability of tests at that time to individuals in the general population who were not exhibiting COVID-19-related symptoms.

**Table 1.** COVID-19 outcome definitions from the Tufts Equity in Health, Wealth, and Civic Engagement Study (2020-2022)

| **Variable** | **Criteria to be classified affirmatively** |
| --- | --- |
| Willingness to vaccinate against COVID-19 | <u>Wave 1</u><br>Response of “yes” to: *“If a vaccine became available to prevent the Coronavirus, would you get it?”*<br><u>Wave 2</u><br>Response of “yes” or “very likely” to:<br>• *“Have you received a COVID-19 vaccine?”*<br>• *“How likely is it that you will get the vaccine when you are eligible?”*<br><u>Wave 3</u><br>Response of “1,” “2,” “3,” or “4” to: *How many doses of a COVID-19 vaccine have you received?* |
| COVID-19 health impact | <u>Wave 1</u><br>Response of “yes” to any of:<br>• *“Have you received a test for the Coronavirus?”*<br>• *“Has anyone in your family received a test for the virus?”*<br>• *“Do you believe you have been personally infected with Coronavirus?”*<br>• *“Have you personally been told by a healthcare professional that you were infected with Coronavirus?”*<br>• *“Has someone in your family or household been told by a healthcare professional that s/he was infected with Coronavirus?”*<br><u>Wave 2</u><br>Response of “yes” to any of:<br>• *“Have you ever tested positive for COVID-19?”*<br>• *“Although you did not receive a positive test for COVID-19, do you believe you have ever had COVID-19?”*<br><u>Wave 3</u><br>Response of “yes” to any of:<br>• *“Do you believe that you have had COVID-19?”*<br>• *“Have you delayed or avoided medical care due to concerns related to COVID-19?”* (i.e., responses of yes to delaying any of emergency care, urgent care, or routine care) |
| COVID-19 assistance recipient | <u>Wave 1</u><br>Response of “yes” to any of:<br>• *“Has the government helped you deal with the Coronavirus or its effects?”*<br>• *“Has any local nonprofit organization or group helped you deal with the Coronavirus or its effects?”*<br>• *“Have individuals from outside your own household helped you deal with the Coronavirus or its effects?”*<br><u>Wave 2</u><br>Response of “yes” to any of:<br>• *“Has the government helped you deal with COVID-19 or its effects?”*<br>• *“Has any local nonprofit organization or group helped you deal with COVID-19 or its effects?”* |
| COVID-19 assistance provider | <u>Waves 1 and 2</u><br>Response of “yes” to either:<br>• *Have you volunteered your time to help other people or organizations* |
|  | *deal with Coronavirus [wave 2: COVID-19]?”*<br>• *“Have you provided health care to anyone else who has had Coronavirus [wave 2: COVID-19]?”* |

### Environmental exposure data

We accounted for four county-level environmental context exposures (air pollution, greenness, toxic release inventory sites, and heatwaves) assessed prior to the first wave of the Tufts Equity in Health, Wealth, and Civic Engagement Study. Based on participants’ county of residence in each of survey waves 1 and 2, we assessed 2018 annual average fine particulate matter (PM_2.5_; particles <2.5 µm in aerodynamic diameter) exposure using North American-specific publicly available models form the University of Washington in Saint Louis Atmospheric Composition Analysis Group (https://sites.wustl.edu/acag/datasets/surface-pm2-5/). The models were derived from combined satellite (aerosol optical depth; Terra and Aqua satellites) and ground-monitoring data.^32, 33^ We created a dichotomous exposure variable for greenness based on summer-time average 16-day composites of normalized difference vegetation index (NDVI; an index that indicates photosynthetic activity in plants; values between −1 [indicating water] and 1 [indicating dense green forests]) using the Moderate Resolution Imaging Spectroradiometer (MODIS) sensor at 250–m x 250–m resolution onboard the Terra satellite (the mean across the county of pixel-level non-negative maximum NDVI from April-September 2018).^34^ Participants were considered to live in areas with low NDVI values if their assigned value was ≤0.6.^35^ To characterize potential residential exposure to toxic pollution, we used location data from the US Environmental Protection Agency’s Toxics Release Inventory (TRI), which provides the locations of facilities that pollute certain regulated chemicals known to have harmful effects on human health and/or the environment.^36^ We created a dichotomous variable representing residence in a county with ≥7 TRI sites based on 2018 data. For example, a county with 30 TRI sites within its borders would be coded as a “1” but a county with only six sites would be coded as “0.” To assess heatwave exposure, we first determined the number of times in 2018 during the warm season (May-September) when the maximum temperature exceeded the pixel-specific 95^th^ percentile for the warm season maximum temperature for 1999-2018 for two or more consecutive days. We then averaged pixel values across tracts and counties (rounding to the nearest whole number) to determine the number of heatwave days per county. We created a dichotomous variable indicating if there were at least three instances of heatwave days.

### Other population-level covariates

We determined two additional ZIP code-level covariates using American Community Survey 5-year (2016-2020) estimates. Both variables were linked to participants’ residential ZIP code at the earliest study wave for which we had this information. One variable was annual household income in 2020 inflation-adjusted dollars.^37^ The other was a proxy of residential racial segregation (derived using B03002 from the US Census Bureau).^38^ Residential racial segregation can take on values between −1 and 1 where values of 0 represent ZIP codes with the same number of people who identify as non-Hispanic Black and non-Hispanic White, values of −1 represent ZIP codes with only people who identify as non-Hispanic Black and none who identify as non-Hispanic White, and values of +1 represent ZIP codes with only people who identify as non-Hispanic White and none who identify as non-Hispanic Black. We categorized this variable to represent racially segregated and mostly non-Hispanic Black ZIP codes (values ≤-0.5), racially heterogeneous ZIP codes (values >-0.5 and <0.5), and racially segregated and mostly non-Hispanic White ZIP codes (values ≥0.5). Similar methods have been used to show disparities in previous epidemiological analyses.^39, 40^

### Statistical analyses

All analyses were conducted in Stata SE v17 using survey weights to maximize representativeness of the sample. We first produced the weighted count and proportion (or mean and 95% confidence interval [CI]) for each exposure, outcome, and covariate for all participants at wave 1 (n = 1267), all participants at wave 2 (n = 1449), and all wave 3 participants who also participated in at least one earlier survey wave (n = 1071). We then fit four sets of multivariable logistic regression models for each exposure-outcome relationship using (1) exposures and outcomes assessed at wave 1, (2) exposures and outcomes assessed at wave 2, (3) exposures assessed at wave 1 and outcomes assessed at wave 2, and (4) exposures assessed at the earliest available time between wave 1 and 2 and outcomes assessed at wave 3. For each set of models we produced five sets of estimates: (1) unadjusted; (2) adjusted for age, gender, educational attainment, and race/ethnicity [primary models]; (3) adjusted for model 2 covariates and discrimination experience, residence in a metropolitan statistical area, ZIP-code level household median income, and ZIP-code level residential racial segregation; (4) stratified by race/ethnicity (non-Hispanic White or any race/ethnicity other than non-Hispanic White) and adjusted for age, gender, and educational attainment; and (5) stratified by discrimination experience (yes/no at the survey wave when exposures were assessed) and adjusted for age, gender, and educational attainment. We also calculated interaction terms for each exposure-stratification variable for each model fit in sets four and five (using multiplicative interaction terms instead of stratification). Additionally, for each set of models with climate stress as the environmental exposure and vaccination as the outcome, we ran sensitivity analyses additionally adjusting for political affiliation.

Finally, we examined associations between a summary environmental context latent variable and each COVID-19 outcome. This analysis is summarized conceptually in Figure 2; notably, our environmental context latent variable should be cautiously interpreted as a preliminary approach to jointly consider the relationship between multiple correlated environmental stressors and COVID-19 outcomes. Analytically, we estimated generalized structural equation models including a latent exposure variable for environmental context (each measured exposure serves as an indicator for the latent variable, which captures the interrelationships among these environmental variables), measured covariates (age, gender, dichotomized educational attainment [greater than high school or not], and dichotomized race/ethnicity [non-Hispanic White or not]), and a measured outcome variable (each outcome modeled separately). All continuous variables were modeled as Gaussian with an identity link function. All dichotomous variables were modeled as Bernoulli distributions with a logit link function. We used the default integration method (mean-variance adaptive Gauss–Hermite quadrature) and constant-only starting values. We examined the direct effect of the latent environmental context variable on the outcome variable using the same four time point sets for exposure and outcome assessment as in the primary regression models.

**Figure 2.**
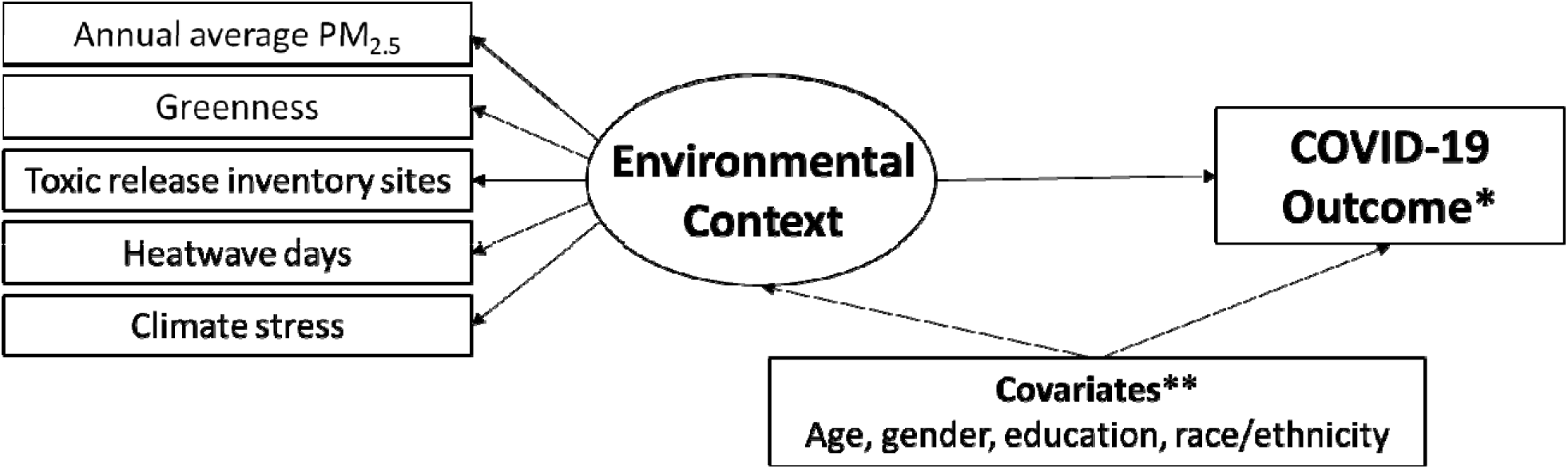
Conceptual model of the relationship between a preliminary environmental context latent construct and COVID-19 outcomes. The latent construct variable (environmental context) is in the circle, and is represented by the measured environmental indicator variables. *In each model, a single COVID-19 outcome variable was assessed (vaccination willingness, health impact, assistance recipient, assistance provider). **In each model, each covariate was included as a predictor of each environmental indicator and the COVID-19 outcome.

## Results

Sample characteristics for all participants in wave 1, all participants in wave 2, and all participants in wave 3 who also participated in an earlier survey wave are shown in Table 2. Based on participants’ wave 1 addresses, annual average PM_2.5_ was 7.4 µg/m^3^, 22% were exposed to low levels of greenness, 79% were exposed to high numbers of toxic release inventory sites, 73% experienced high numbers of heatwave days, and 53% experienced climate stress. Annual average PM_2.5_ was significantly associated with toxic release inventory sites and heatwave days (waves 1 and 2), heatwave days were significantly associated with greenness and toxic release inventory sites (waves 1 and 2), and greenness was significantly associated with toxic release inventory sites (wave 2). In survey wave 1 (spring 2020), 58% of participants indicated that they were willing to get vaccinated, 26% had personally experienced or had a family member experience a COVID-19 health impact, 10% had provided assistance for COVID-19, and 41% had received assistance for COVID-19. In survey wave 3 (spring 2022), 80% of participants had reported receiving at least one vaccination against COVID-19 and 46% had a COVID-19 health impact (had COVID-19 and/or delayed medical care due to COVID-19).

**Table 2.** Sample characteristics of the Tufts Equity in Health, Wealth, and Civic Engagement Study participants

|  | **Characteristics at wave 1 (2020) for wave 1 participants**<br>Weighted % (weighted n) or weighted mean (95% CI)<br>n = 1267 | **Characteristics at wave 2 (2021) for wave 2 participants**<br>Weighted % (weighted n) or weighted mean (95% CI)<br>n = 1449 | **Characteristics at the earliest available wave of wave 1 or 2 for wave 3 participants**<br>Weighted % (weighted n) or weighted mean (95% CI)<br>n = 1071 |
| --- | --- | --- | --- |
| **Age (years)** | 48.0 (46.9, 49.1) | 48.4 (47.2, 49.6) | 47.4 (45.9, 48.8) |
| **Gender** |  |  |  |
| Female | 51.6 (654) | 51.6 (748) | 51.2 (548) |
| Male | 48.4 (613) | 48.4 (701) | 48.8 (523) |
| **Income** |  |  |  |
| <\$25K | 13.6 (172) | 12.6 (182) | 12.7 (137) |
| \$25K-<\$50K | 18.2 (231) | 17.6 (255) | 15.5 (166) |
| \$50K-<\$75K | 17.2 (218) | 17.4 (252) | 16.3 (175) |
| \$75K-<\$100K | 13.7 (174) | 14.0 (203) | 13.0 (139) |
| \$100K-<\$150K | 17.7 (224) | 18.5 (269) | 19.7 (211) |
| ≥\$150K | 19.6 (248) | 19.9 (288) | 22.7 (244) |
| **Education** |  |  |  |
| Less than high school | 10.6 (134) | 11.2 (162) | 10.4 (112) |
| High school diploma or equivalent | 28.3 (359) | 27.4 (396) | 30.8 (330) |
| Some college or Associate's | 27.8 (352) | 29.9 (433) | 26.9 (288) |
| Bachelor's degree or higher | 33.3 (422) | 31.6 (457) | 31.9 (341) |
| **Race/ethnicity** |  |  |  |
| Non-Hispanic White | 63.1 (800) | 62.9 (912) | 62.7 (672) |
| Non-Hispanic Black/African American | 11.8 (150) | 11.8 (171) | 12.2 (131) |
| Non-Hispanic other or multiracial | 8.6 (109) | 8.7 (126) | 8.2 (88) |
| Hispanic | 16.4 (208) | 16.5 (240) | 16.8 (180) |
| **Discrimination experience<sup>1</sup>** |  |  |  |
| No | 40.0 (503) | 43.9 (627) | 44.5 (472) |
| Yes | 60.0 (753) | 56.1 (801) | 55.5 (589) |
| **Residence in a metropolitan area** |  |  |  |
| No | 13.3 (169) | 13.4 (195) | 13.7 (147) |
| Yes | 86.7 (1098) | 86.6 (1254) | 86.3 (924) |
| **Racial segregation of ZIP-code** |  |  |  |
| Segregated, mostly non-Hispanic White | 82.1 (616) | 74.6 (735) | 75.9 (548) |
| Less segregated | 13.4 (101) | 19.2 (189) | 18.9 (137) |
| Segregated, mostly non-Hispanic Black | 4.5 (33) | 6.3 (62) | 5.2 (38) |
| **ZIP-code level median household income (\$)** | 71647 (69158, 74135) | 69008 (66745, 71272) | 68437 (65864, 71010) |
| **Exposures<sup>2</sup>** |  |  |  |
| Annual average PM<sub>2.5</sub> (µg/m<sup>3</sup>) | 7.4 (7.3, 7.5) | 7.5 (7.4, 7.6) | 7.3 (7.2, 7.4) |
| Low greenness | 21.6 (274) | 21.3 (308) | 21.4 (229) |
| High toxic release inventory | 79.3 (1003) | 79.9 (1157) | 79.4 (850) |
| High heatwave day | 72.7 (912) | 71.5 (1027) | 73.1 (775) |
| High climate stress | 52.8 (662) | 38.2 (545) | 49.4 (524) |
| **Outcomes<sup>3</sup> (at wave 1 for column 2, at wave 2 for columns 3 and 4)** |  |  |  |
| Willingness to vaccinate | 57.8 (726) | 66.1 (949) | 67.2 (672) |
| COVID-19 health impact | 25.8 (322) | 10.4 (146) | 9.7 (96) |
| Receipt of COVID-19 | 41.4 (516) | 24.1 (346) | 25.5 (255) |
| assistance |  |  |  |
| Provision of COVID-19 assistance | 10.3 (128) | 10.4 (149) | 10.8 (109) |

<sup>1</sup>Discrimination experience based on affirmative answer to any question pertaining to perceived discrimination in everyday situations (e.g., receiving poorer service than others in restaurants and stores; you have been unfairly stopped, searched, questioned, physically threatened, or abused by the police).
<sup>2</sup>Environmental variables include county-level annual air pollution exposure, greenness exposure (low = normalized difference vegetation index $\leq 0.6$ ), Toxics Release Inventory sites exposure (high = residence in a county with $\geq 7$ toxic release inventory sites), heatwave day exposure (high = residence in a county with $\geq 3$ instances of heatwave days in 2018), and individual-level climate stress (high = affirmative response that climate change/global warming is stressful/makes the participant anxious).
<sup>3</sup>Outcome variables have slightly different definitions at each survey cycle (see Table 1).

Several environmental exposures were significantly associated with willingness to be vaccinated against COVID-19 (Table 3). For example, residence in wave 1 in an area with more TRI sites was associated with higher likelihood of being willing to vaccinate in wave 1. Additionally, low greenness exposure in wave 2 and high heatwave day exposure in wave 2 were each positively associated with being willing to vaccinate in wave 2. Overall, the most robust association with vaccination willingness was climate stress (agreement with climate stress was associated with increased likelihood of vaccination; p < 0.01 in every model regardless of survey wave for exposures or outcomes and regardless of covariate choice). However, this association between climate stress and vaccination willingness was attenuated (and not significant in some models) for participants who identified as any race/ethnicity other than non-Hispanic White (Table 3). Furthermore, when adjusting for political affiliation, some of the associations were attenuated, though the cross-sectional wave 2 associations were robust (Supplemental Table 1).

**Table 3.** Associations between environmental exposures^1^ and self-reported willingness to vaccinate from the 2020-2022 Tufts Equity in Health, Wealth, and Civic Engagement Study

|  | **Exposure: W1<sup>2</sup>**<br>**Outcome: W1**<br>OR (95% CI); p | **Exposure: W2**<br>**Outcome: W2**<br>OR (95% CI); p | **Exposure: W1**<br>**Outcome: W2**<br>OR (95% CI); p | **Exposure: W1-2<sup>3</sup>**<br>**Outcome: W3**<br>OR (95% CI); p |
| --- | --- | --- | --- | --- |
| **Annual average PM<sub>2.5</sub> (µg/m<sup>3</sup>)** |  |  |  |  |
| Unadjusted | 0.99 (0.92, 1.06);<br>0.736 | 0.98 (0.90, 1.08);<br>0.720 | 1.02 (0.91, 1.14);<br>0.737 | 1.05 (0.94, 1.18);<br>0.367 |
| Adjusted 1<sup>4</sup> | 0.99 (0.92, 1.07);<br>0.767 | 1.01 (0.92, 1.10);<br>0.873 | 1.05 (0.94, 1.18);<br>0.357 | 1.01 (0.89, 1.16);<br>0.833 |
| Adjusted 2<sup>5</sup> | 0.96 (0.86, 1.06);<br>0.405 | 0.97 (0.87, 1.07);<br>0.501 | 1.02 (0.88, 1.18);<br>0.839 | 0.97 (0.83, 1.13);<br>0.682 |
| Non-Hispanic White<sup>6</sup> | 0.99 (0.90, 1.08);<br>0.810 | 0.96 (0.85, 1.09);<br>0.558 | 0.96 (0.85, 1.10);<br>0.578 | 1.01 (0.83, 1.24);<br>0.923 |
| Other than non-Hispanic White | 0.97 (0.84, 1.12);<br>0.669 | 1.08 (0.96, 1.21);<br>0.185 | 1.18 (0.96, 1.46);<br>0.114 | 1.02 (0.86, 1.21);<br>0.790 |
| No discrimination<sup>7</sup> | 1.00 (0.88, 1.13);<br>0.978 | 1.14 (0.99, 1.31);<br>0.065 | 1.12 (0.92, 1.37);<br>0.258 | 0.99 (0.83, 1.18);<br>0.919 |
| Discrimination | 0.99 (0.90, 1.09);<br>0.828 | 0.94 (0.84, 1.06);<br>0.318 | 1.05 (0.92, 1.20);<br>0.446 | 1.16 (0.97, 1.39);<br>0.111 |
| **Greenness** |  |  |  |  |
| Unadjusted | 1.15 (0.84, 1.56);<br>0.389 | **1.64 (1.19, 2.25);**<br>**0.002\*** | 1.39 (0.89, 2.19);<br>0.148 | **1.85 (1.14, 3.01);**<br>**0.013\*** |
| Adjusted 1 | 1.04 (0.74, 1.46);<br>0.822 | **1.45 (1.01, 2.08);**<br>**0.045\*** | 0.92 (0.56, 1.51);<br>0.753 | 1.45 (0.82, 2.58);<br>0.201 |
| Adjusted 2 | 0.88 (0.58, 1.34);<br>0.552 | 1.47 (0.97, 2.22);<br>0.070 | 0.89 (0.49, 1.63);<br>0.705 | 1.21 (0.64, 2.29);<br>0.547 |
| Non-Hispanic White | 1.45 (0.94, 2.23);<br>0.095 | 1.05 (0.59, 1.87);<br>0.880 | 1.11 (0.62, 2.00);<br>0.725 | 0.78 (0.37, 1.65);<br>0.512 |
| Other than non-Hispanic White | 0.89 (0.53, 1.47);<br>0.637 | **1.93 (1.29, 2.89);**<br>**0.002\*** | 1.16 (0.57, 2.35);<br>0.684 | **2.18 (1.03, 4.61);**<br>**0.042\*** |
| No discrimination | 1.13 (0.67, 1.89);<br>0.656 | 1.59 (0.94, 2.70);<br>0.086 | 1.17 (0.52, 2.63);<br>0.702 | 2.22 (0.93, 5.27);<br>0.072 |
| Discrimination | 1.19 (0.77, 1.83);<br>0.437 | **1.79 (1.13, 2.84);**<br>**0.013\*** | 1.78 (0.94, 3.38);<br>0.078 | **2.27 (1.08, 4.79);**<br>**0.032\*** |
| **Toxic release inventory sites** |  |  |  |  |
| Unadjusted | **1.47 (1.09, 1.98);**<br>**0.012\*** | 1.13 (0.80, 1.60);<br>0.495 | 1.33 (0.89, 2.00);<br>0.165 | 1.44 (0.88, 2.34);<br>0.145 |
| Adjusted 1 | **1.39 (1.01, 1.90);**<br>**0.042\*** | 1.17 (0.81, 1.69);<br>0.389 | 1.29 (0.80, 2.06);<br>0.296 | 1.39 (0.88, 2.21);<br>0.159 |
| Adjusted 2 | **1.68 (1.07, 2.63);**<br>**0.023\*** | 0.79 (0.49, 1.28);<br>0.336 | 0.95 (0.49, 1.83);<br>0.880 | 0.72 (0.38, 1.38);<br>0.320 |
| Non-Hispanic White | 1.38 (0.96, 1.99);<br>0.083 | 1.54 (0.92, 2.58);<br>0.098 | 1.57 (0.93, 2.66);<br>0.094 | **1.82 (1.05, 3.15);**<br>**0.034\*** |
| Other than non-Hispanic White | 1.44 (0.76, 2.74);<br>0.264 | 0.73 (0.45, 1.18);<br>0.201 | 0.90 (0.36, 2.28);<br>0.827 | 0.92 (0.35, 2.40);<br>0.870 |
| No discrimination | **1.73 (1.08, 2.75);**<br>**0.022\*** | **1.78 (1.06, 2.97);**<br>**0.028\*** | 1.84 (0.94, 3.60);<br>0.074 | 1.49 (0.72, 3.07);<br>0.282 |
| Discrimination | 1.20 (0.79, 1.83);<br>0.400 | 0.92 (0.54, 1.54);<br>0.742 | 1.04 (0.55, 1.98);<br>0.905 | 1.41 (0.74, 2.71);<br>0.296 |
| **Heatwave days** |  |  |  |  |
| Unadjusted | 1.11 (0.83, 1.47);<br>0.486 | **1.45 (1.06, 1.98);**<br>**0.021\*** | **1.51 (1.03, 2.20);**<br>**0.033\*** | 1.46 (0.92, 2.33);<br>0.112 |
| Adjusted 1 | 1.07 (0.79, 1.44);<br>0.674 | **1.39 (1.00, 1.91);**<br>**0.047\*** | 1.39 (0.91, 2.13);<br>0.127 | 1.47 (0.90, 2.38);<br>0.122 |
| Adjusted 2 | 0.93 (0.61, 1.43);<br>0.751 | 1.38 (0.93, 2.03);<br>0.109 | 1.17 (0.65, 2.12);<br>0.602 | 1.16 (0.62, 2.17);<br>0.649 |
| Non-Hispanic White | **1.48 (1.05, 2.09);**<br>**0.024\*** | 1.45 (0.91, 2.31);<br>0.120 | **1.63 (1.01, 2.63);**<br>**0.046\*** | 1.59 (0.86, 2.96);<br>0.141 |
| Other than non-Hispanic White | 0.68 (0.39, 1.19);<br>0.172 | 1.46 (0.98, 2.17);<br>0.064 | 1.55 (0.73, 3.29);<br>0.256 | 1.31 (0.63, 2.72);<br>0.475 |
| No discrimination | 1.49 (0.96, 2.31);<br>0.076 | **1.66 (1.02, 2.69);**<br>**0.041\*** | **1.90 (1.03, 3.49);**<br>**0.040\*** | **2.39 (1.21, 4.72);**<br>**0.012\*** |
| Discrimination | 0.95 (0.64, 1.42);<br>0.809 | 1.36 (0.89, 2.07);<br>0.150 | 1.59 (0.92, 2.75);<br>0.094 | 0.96 (0.49, 1.88);<br>0.895 |
| **Climate stress** |  |  |  |  |
| Unadjusted | **2.50 (1.93, 3.24);**<br>**<0.001\*** | **4.52 (3.22, 6.33);**<br>**<0.001\*** | **2.53 (1.78, 3.62);**<br>**<0.001\*** | **2.50 (1.58, 3.97);**<br>**<0.001\*** |
| Adjusted 1 | **2.77 (2.09, 3.69);**<br>**<0.001\*** | **4.27 (2.99, 6.11);**<br>**<0.001\*** | **2.72 (1.80, 4.11);**<br>**<0.001\*** | **2.35 (1.47, 3.76);**<br>**<0.001\*** |
| Adjusted 2 | **3.32 (2.26, 4.89);**<br>**<0.001\*** | **4.42 (2.93, 6.65);**<br>**<0.001\*** | **6.15 (3.29, 11.47);**<br>**<0.001\*** | **2.69 (1.45, 5.00);**<br>**0.002\*** |
| Non-Hispanic White | **3.76 (2.68, 5.29);**<br>**<0.001\*** | **6.09 (3.37, 11.01);**<br>**<0.001\*** | **3.73 (2.31, 6.01);**<br>**<0.001\*** | **3.37 (1.84, 6.17);**<br>**<0.001\*** |
| Other than non-Hispanic White | 1.62 (0.97, 2.70);<br>0.064 | **3.05 (2.00, 4.64);**<br>**<0.001\*** | 1.68 (0.84, 3.39);<br>0.143 | 1.86 (0.86, 4.00);<br>0.114 |
| No discrimination | 4.48 (2.86, 7.02);<br><0.001* | 6.88 (3.71, 12.76);<br><0.001* | 5.42 (2.59, 11.32);<br><0.001* | 2.83 (1.30, 6.18);<br>0.009* |
| Discrimination | 2.28 (1.56, 3.33);<br><0.001* | 3.78 (2.44, 5.87);<br><0.001* | 2.54 (1.47, 4.39);<br><0.001* | 2.36 (1.28, 4.36);<br>0.006* |

<sup>1</sup>Environmental variables include county-level annual air pollution exposure, greenness exposure (low = normalized difference vegetation index $\leq 0.6$ ), toxic release inventory exposure (high = residence in a county with $\geq 7$ toxic release inventory sites), heatwave day exposure (high = residence in a county with $\geq 3$ instances of heatwave days in 2018), and individual-level climate stress (high = affirmative response that climate change/global warming is stressful/makes the participant anxious).
<sup>2</sup>W1 = wave 1; W2 = wave 2; W3 = wave 3. Outcome definition varied by survey wave (see Table 1).
<sup>3</sup>W1-2 = Defined by their value at the earliest survey wave for which participants had data.
<sup>4</sup>Adjusted for age, gender, educational attainment, and race/ethnicity [primary models].
<sup>5</sup>Adjusted for primary model covariates and discrimination experience, residence in a metropolitan region, ZIP-code level household median income, and ZIP-code level residential racial segregation.
<sup>6</sup>Stratified by race/ethnicity (non-Hispanic White or any race/ethnicity other than non-Hispanic White) and adjusted for age, gender, and educational attainment.
<sup>7</sup>Stratified by perceived discrimination experience (yes/no) and adjusted for age, gender, and educational attainment.
\*p < 0.05

No environmental exposure variable was associated with COVID-19 health impacts in the unadjusted or adjusted models (Supplemental Table 2). The only exception was for the association between TRI assessed in wave 2 and COVID-19 health impacts assessed in wave 2 among people who identify as a race/ethnicity other than non-Hispanic White (OR = 0.41; 95% CI = 0.22, 0.75) and among people who reported experiences of discrimination in wave 2 (OR = 0.50; 95% CI = 0.26, 0.96). There was also evidence of a statistically significant interaction between climate stress and discrimination experience among participants in the second wave of the survey (p = 0.045; inverse association among those without experience of discrimination but positive association among those with experience of discrimination; Supplemental Tables 2 and 3).

Among the environmental variables, only climate stress was significantly associated with receipt of assistance for COVID-19 in the unstratified models (Table 4). This relationship was most robust between climate stress reported in wave 2 and COVID-19 outcomes reported in wave 2 (Table 4). Low greenness exposure in wave 1 was associated with higher likelihood of COVID-19 assistance receipt in wave 1 among people who identified as any race/ethnicity other than non-Hispanic White (OR = 1.66, 95% CI = 1.02, 2.71). Similarly, more heatwave days exposure in wave 2 was associated with higher likelihood of COVID-19 assistance receipt in wave 2 among people who identified as any race/ethnicity other than non-Hispanic White (OR = 1.88, 95% CI = 1.13, 3.11) and among people who experienced discrimination (OR = 1.78, 95% CI = 1.12, 2.82). Discrimination experience at wave 1 interacted with air pollution exposure at wave 2 to affect the likelihood of COVID-19 assistance receipt in wave 2 (p for interaction = 0.024; significant inverse association among those without discrimination experience; Table 4 and Supplemental Table 3).

**Table 4.**
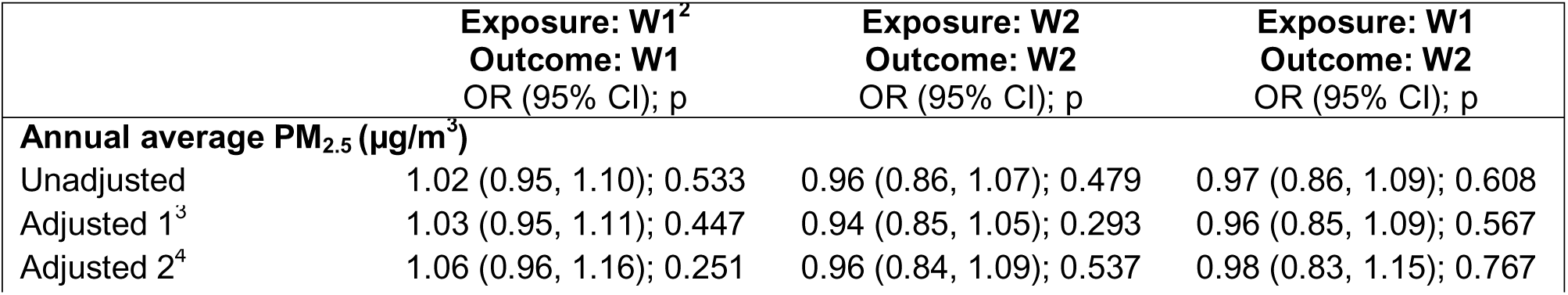
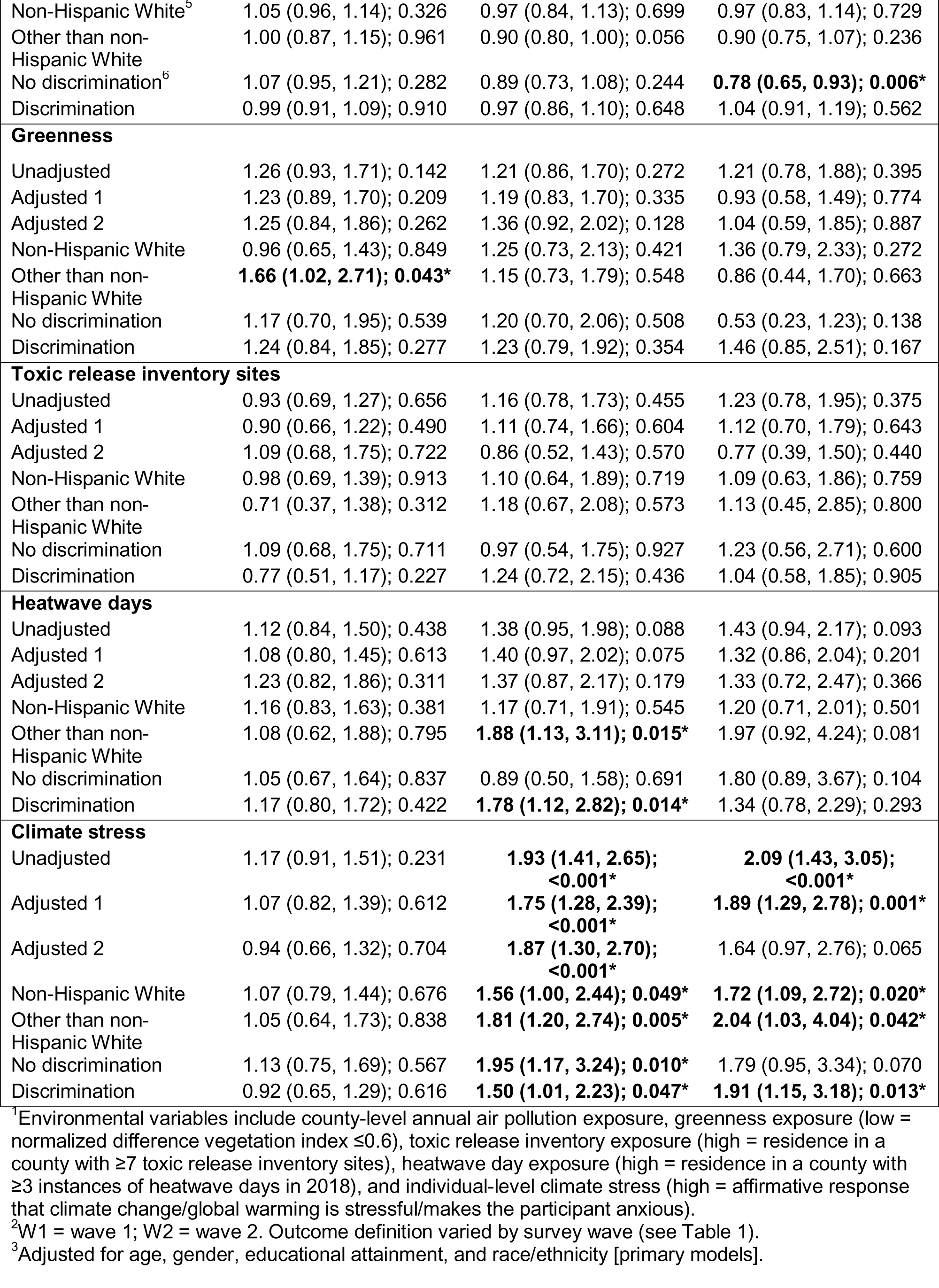
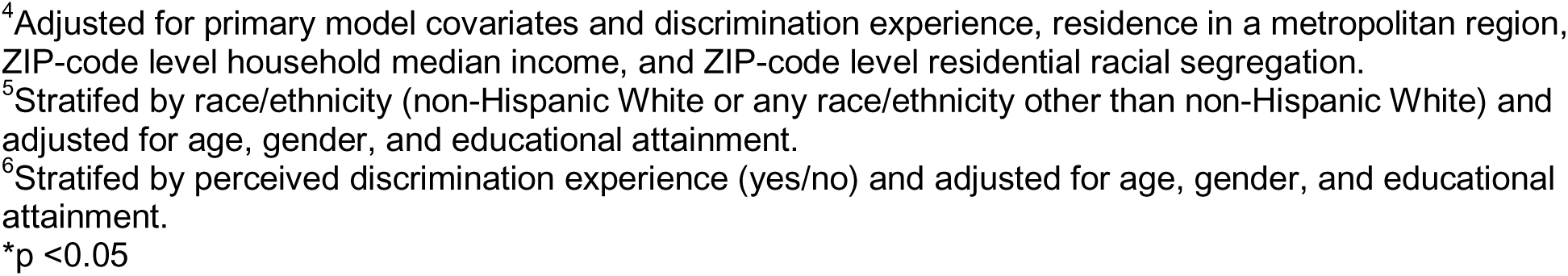
Associations between environmental exposures^1^ and self-reported receipt of COVID-19 assistance from the 2020-2022 Tufts Equity in Health, Wealth, and Civic Engagement Study

|  | Exposure: W1<sup>2</sup><br>Outcome: W1<br>OR (95% CI); p | Exposure: W2<br>Outcome: W2<br>OR (95% CI); p | Exposure: W1<br>Outcome: W2<br>OR (95% CI); p |
| --- | --- | --- | --- |
| **Annual average PM<sub>2.5</sub> (µg/m<sup>3</sup>)** |  |  |  |
| Unadjusted | 1.02 (0.95, 1.10); 0.533 | 0.96 (0.86, 1.07); 0.479 | 0.97 (0.86, 1.09); 0.608 |
| Adjusted 1<sup>3</sup> | 1.03 (0.95, 1.11); 0.447 | 0.94 (0.85, 1.05); 0.293 | 0.96 (0.85, 1.09); 0.567 |
| Adjusted 2<sup>4</sup> | 1.06 (0.96, 1.16); 0.251 | 0.96 (0.84, 1.09); 0.537 | 0.98 (0.83, 1.15); 0.767 |
| Non-Hispanic White<sup>5</sup> | 1.05 (0.96, 1.14); 0.326 | 0.97 (0.84, 1.13); 0.699 | 0.97 (0.83, 1.14); 0.729 |
| Other than non-Hispanic White | 1.00 (0.87, 1.15); 0.961 | 0.90 (0.80, 1.00); 0.056 | 0.90 (0.75, 1.07); 0.236 |
| No discrimination<sup>6</sup> | 1.07 (0.95, 1.21); 0.282 | 0.89 (0.73, 1.08); 0.244 | **0.78 (0.65, 0.93); 0.006\*** |
| Discrimination | 0.99 (0.91, 1.09); 0.910 | 0.97 (0.86, 1.10); 0.648 | 1.04 (0.91, 1.19); 0.562 |
| **Greenness** |  |  |  |
| Unadjusted | 1.26 (0.93, 1.71); 0.142 | 1.21 (0.86, 1.70); 0.272 | 1.21 (0.78, 1.88); 0.395 |
| Adjusted 1 | 1.23 (0.89, 1.70); 0.209 | 1.19 (0.83, 1.70); 0.335 | 0.93 (0.58, 1.49); 0.774 |
| Adjusted 2 | 1.25 (0.84, 1.86); 0.262 | 1.36 (0.92, 2.02); 0.128 | 1.04 (0.59, 1.85); 0.887 |
| Non-Hispanic White | 0.96 (0.65, 1.43); 0.849 | 1.25 (0.73, 2.13); 0.421 | 1.36 (0.79, 2.33); 0.272 |
| Other than non-Hispanic White | **1.66 (1.02, 2.71); 0.043\*** | 1.15 (0.73, 1.79); 0.548 | 0.86 (0.44, 1.70); 0.663 |
| No discrimination | 1.17 (0.70, 1.95); 0.539 | 1.20 (0.70, 2.06); 0.508 | 0.53 (0.23, 1.23); 0.138 |
| Discrimination | 1.24 (0.84, 1.85); 0.277 | 1.23 (0.79, 1.92); 0.354 | 1.46 (0.85, 2.51); 0.167 |
| **Toxic release inventory sites** |  |  |  |
| Unadjusted | 0.93 (0.69, 1.27); 0.656 | 1.16 (0.78, 1.73); 0.455 | 1.23 (0.78, 1.95); 0.375 |
| Adjusted 1 | 0.90 (0.66, 1.22); 0.490 | 1.11 (0.74, 1.66); 0.604 | 1.12 (0.70, 1.79); 0.643 |
| Adjusted 2 | 1.09 (0.68, 1.75); 0.722 | 0.86 (0.52, 1.43); 0.570 | 0.77 (0.39, 1.50); 0.440 |
| Non-Hispanic White | 0.98 (0.69, 1.39); 0.913 | 1.10 (0.64, 1.89); 0.719 | 1.09 (0.63, 1.86); 0.759 |
| Other than non-Hispanic White | 0.71 (0.37, 1.38); 0.312 | 1.18 (0.67, 2.08); 0.573 | 1.13 (0.45, 2.85); 0.800 |
| No discrimination | 1.09 (0.68, 1.75); 0.711 | 0.97 (0.54, 1.75); 0.927 | 1.23 (0.56, 2.71); 0.600 |
| Discrimination | 0.77 (0.51, 1.17); 0.227 | 1.24 (0.72, 2.15); 0.436 | 1.04 (0.58, 1.85); 0.905 |
| **Heatwave days** |  |  |  |
| Unadjusted | 1.12 (0.84, 1.50); 0.438 | 1.38 (0.95, 1.98); 0.088 | 1.43 (0.94, 2.17); 0.093 |
| Adjusted 1 | 1.08 (0.80, 1.45); 0.613 | 1.40 (0.97, 2.02); 0.075 | 1.32 (0.86, 2.04); 0.201 |
| Adjusted 2 | 1.23 (0.82, 1.86); 0.311 | 1.37 (0.87, 2.17); 0.179 | 1.33 (0.72, 2.47); 0.366 |
| Non-Hispanic White | 1.16 (0.83, 1.63); 0.381 | 1.17 (0.71, 1.91); 0.545 | 1.20 (0.71, 2.01); 0.501 |
| Other than non-Hispanic White | 1.08 (0.62, 1.88); 0.795 | **1.88 (1.13, 3.11); 0.015\*** | 1.97 (0.92, 4.24); 0.081 |
| No discrimination | 1.05 (0.67, 1.64); 0.837 | 0.89 (0.50, 1.58); 0.691 | 1.80 (0.89, 3.67); 0.104 |
| Discrimination | 1.17 (0.80, 1.72); 0.422 | **1.78 (1.12, 2.82); 0.014\*** | 1.34 (0.78, 2.29); 0.293 |
| **Climate stress** |  |  |  |
| Unadjusted | 1.17 (0.91, 1.51); 0.231 | **1.93 (1.41, 2.65); <0.001\*** | **2.09 (1.43, 3.05); <0.001\*** |
| Adjusted 1 | 1.07 (0.82, 1.39); 0.612 | **1.75 (1.28, 2.39); <0.001\*** | **1.89 (1.29, 2.78); 0.001\*** |
| Adjusted 2 | 0.94 (0.66, 1.32); 0.704 | **1.87 (1.30, 2.70); <0.001\*** | 1.64 (0.97, 2.76); 0.065 |
| Non-Hispanic White | 1.07 (0.79, 1.44); 0.676 | **1.56 (1.00, 2.44); 0.049\*** | **1.72 (1.09, 2.72); 0.020\*** |
| Other than non-Hispanic White | 1.05 (0.64, 1.73); 0.838 | **1.81 (1.20, 2.74); 0.005\*** | **2.04 (1.03, 4.04); 0.042\*** |
| No discrimination | 1.13 (0.75, 1.69); 0.567 | **1.95 (1.17, 3.24); 0.010\*** | 1.79 (0.95, 3.34); 0.070 |
| Discrimination | 0.92 (0.65, 1.29); 0.616 | **1.50 (1.01, 2.23); 0.047\*** | **1.91 (1.15, 3.18); 0.013\*** |

<sup>1</sup>Environmental variables include county-level annual air pollution exposure, greenness exposure (low = normalized difference vegetation index $\leq 0.6$ ), toxic release inventory exposure (high = residence in a county with $\geq 7$ toxic release inventory sites), heatwave day exposure (high = residence in a county with $\geq 3$ instances of heatwave days in 2018), and individual-level climate stress (high = affirmative response that climate change/global warming is stressful/makes the participant anxious).
<sup>2</sup>W1 = wave 1; W2 = wave 2. Outcome definition varied by survey wave (see Table 1).
<sup>3</sup>Adjusted for age, gender, educational attainment, and race/ethnicity [primary models].
<sup>4</sup>Adjusted for primary model covariates and discrimination experience, residence in a metropolitan region, ZIP-code level household median income, and ZIP-code level residential racial segregation.
<sup>5</sup>Stratified by race/ethnicity (non-Hispanic White or any race/ethnicity other than non-Hispanic White) and adjusted for age, gender, and educational attainment.
<sup>6</sup>Stratified by perceived discrimination experience (yes/no) and adjusted for age, gender, and educational attainment.
\*p <0.05

The environmental variable with the most consistent association with provision of assistance for COVID-19 was air pollution exposure. More air pollution exposure in wave 1 was associated with higher likelihood of provisioning COVID-19 assistance in wave 1 (Table 5), especially among people who have experienced discrimination (OR = 1.19, 95% CI = 1.02, 1.39). More TRI exposure in wave 2 was associated with decreased likelihood of provisioning COVID-19 assistance in wave 2, especially among people who identify as any race/ethnicity other than non-Hispanic White (Table 5). Discrimination experience significantly interacted with climate stress in wave 2 to affect likelihood of reporting provisioning assistance in wave 2 (p = 0.013; Supplemental Table 3).

**Table 5.** Associations between environmental exposures^1^ and self-reported provision of COVID-19 assistance from the 2020-2022 Tufts Equity in Health, Wealth, and Civic Engagement Study

|  | **Exposure: W1<sup>2</sup>**<br>**Outcome: W1**<br>OR (95% CI); p | **Exposure: W2**<br>**Outcome: W2**<br>OR (95% CI); p | **Exposure: W1**<br>**Outcome: W2**<br>OR (95% CI); p |
| --- | --- | --- | --- |
| **Annual average PM<sub>2.5</sub> (µg/m<sup>3</sup>)** |  |  |  |
| Unadjusted | **1.16 (1.02, 1.32); 0.021\*** | 0.98 (0.88, 1.09); 0.712 | 1.06 (0.92, 1.22); 0.427 |
| Adjusted 1<sup>3</sup> | **1.16 (1.02, 1.32); 0.026\*** | 0.97 (0.87, 1.09); 0.612 | 1.06 (0.93, 1.22); 0.387 |
| Adjusted 2<sup>4</sup> | 1.14 (0.97, 1.32); 0.103 | 0.94 (0.83, 1.08); 0.390 | 1.05 (0.87, 1.25); 0.621 |
| Non-Hispanic White<sup>5</sup> | 1.12 (0.93, 1.35); 0.244 | 1.01 (0.86, 1.18); 0.933 | 1.02 (0.86, 1.19); 0.850 |
| Other than non-Hispanic White | 1.22 (1.00, 1.48); 0.052 | 0.91 (0.79, 1.06); 0.239 | 1.15 (0.89, 1.47); 0.289 |
| No discrimination<sup>6</sup> | 1.07 (0.88, 1.31); 0.474 | 0.95 (0.81, 1.12); 0.566 | 1.04 (0.85, 1.27); 0.693 |
| Discrimination | **1.19 (1.02, 1.39); 0.030\*** | 1.01 (0.87, 1.17); 0.897 | 1.05 (0.89, 1.24); 0.530 |
| **Greenness** |  |  |  |
| Unadjusted | 1.15 (0.71, 1.84); 0.574 | 1.06 (0.67, 1.68); 0.792 | 1.31 (0.68, 2.52); 0.422 |
| Adjusted 1 | 1.01 (0.62, 1.65); 0.957 | 0.85 (0.52, 1.38); 0.510 | 1.04 (0.52, 2.06); 0.918 |
| Adjusted 2 | 0.88 (0.51, 1.51); 0.631 | 0.95 (0.56, 1.59); 0.835 | 1.24 (0.61, 2.51); 0.547 |
| Non-Hispanic White | 0.71 (0.35, 1.42); 0.328 | 0.63 (0.24, 1.66); 0.347 | 0.57 (0.21, 1.55); 0.272 |
| Other than non-Hispanic White | 1.28 (0.64, 2.54); 0.483 | 1.13 (0.65, 1.95); 0.664 | 1.44 (0.58, 3.53); 0.428 |
| No discrimination | 1.31 (0.54, 3.16); 0.545 | 1.16 (0.58, 2.29); 0.675 | 0.60 (0.19, 1.91); 0.391 |
| Discrimination | 1.08 (0.62, 1.89); 0.790 | 1.00 (0.54, 1.87); 0.994 | 1.66 (0.78, 3.52); 0.187 |
| **Toxic release inventory sites** |  |  |  |
| Unadjusted | 0.97 (0.60, 1.57); 0.895 | 0.65 (0.42, 1.01); 0.057 | 0.63 (0.36, 1.11); 0.112 |
| Adjusted 1 | 0.88 (0.54, 1.44); 0.612 | **0.62 (0.39, 0.97); 0.038\*** | 0.60 (0.34, 1.06); 0.081 |
| Adjusted 2 | 0.92 (0.45, 1.88); 0.828 | 0.94 (0.50, 1.76); 0.848 | 1.04 (0.40, 2.69); 0.930 |
| Non-Hispanic White | 1.22 (0.66, 2.28); 0.524 | 0.71 (0.38, 1.33); 0.278 | 0.72 (0.38, 1.36); 0.309 |
| Other than non-Hispanic White | 0.51 (0.23, 1.14); 0.100 | **0.48 (0.25, 0.89); 0.021\*** | 0.49 (0.17, 1.41); 0.185 |
| No discrimination | 1.44 (0.52, 3.97); 0.484 | 0.53 (0.27, 1.05); 0.069 | 0.85 (0.35, 2.06); 0.723 |
| Discrimination | 0.77 (0.44, 1.36); 0.367 | 0.69 (0.38, 1.25); 0.219 | 0.51 (0.25, 1.05); 0.069 |
| **Heatwave days** |  |  |  |
| Unadjusted | 0.88 (0.55, 1.40); 0.583 | 1.01 (0.65, 1.56); 0.962 | 0.87 (0.52, 1.46); 0.589 |
| Adjusted 1 | 0.86 (0.52, 1.42); 0.551 | 0.94 (0.60, 1.47); 0.784 | 0.77 (0.45, 1.33); 0.346 |
| Adjusted 2 | 0.67 (0.37, 1.21); 0.184 | 1.04 (0.61, 1.79); 0.880 | 0.80 (0.38, 1.68); 0.551 |
| Non-Hispanic White | 1.07 (0.58, 1.97); 0.825 | 0.69 (0.38, 1.25); 0.220 | 0.70 (0.38, 1.27); 0.237 |
| Other than non-Hispanic White | 0.63 (0.29, 1.38); 0.247 | 1.57 (0.83, 2.95); 0.164 | 0.94 (0.34, 2.59); 0.905 |
| No discrimination | 1.24 (0.51, 3.01); 0.640 | 0.86 (0.44, 1.68); 0.656 | 0.59 (0.27, 1.32); 0.202 |
| Discrimination | 0.78 (0.45, 1.37); 0.393 | 1.11 (0.62, 2.01); 0.723 | 1.12 (0.55, 2.27); 0.758 |
| **Climate stress** |  |  |  |
| Unadjusted | 1.40 (0.92, 2.11); 0.113 | 1.22 (0.82, 1.80); 0.322 | 1.37 (0.83, 2.26); 0.224 |
| Adjusted 1 | 1.23 (0.79, 1.92); 0.361 | 1.01 (0.68, 1.51); 0.951 | 1.19 (0.70, 2.01); 0.514 |
| Adjusted 2 | 0.88 (0.49, 1.60); 0.678 | 1.10 (0.69, 1.74); 0.700 | 1.67 (0.78, 3.55); 0.184 |
| Non-Hispanic White | 1.45 (0.85, 2.47); 0.168 | 1.19 (0.67, 2.13); 0.557 | 1.51 (0.85, 2.71); 0.161 |
| Other than non-Hispanic White | 0.87 (0.42, 1.79); 0.704 | 0.84 (0.49, 1.45); 0.528 | 0.88 (0.34, 2.23); 0.781 |
| No discrimination | 1.75 (0.79, 3.87); 0.168 | 0.55 (0.27, 1.10); 0.090 | 0.87 (0.39, 1.97); 0.740 |
| Discrimination | 0.97 (0.58, 1.63); 0.917 | 1.52 (0.91, 2.54); 0.107 | 1.54 (0.75, 3.19); 0.242 |

<sup>1</sup>Environmental variables include county-level annual air pollution exposure, greenness exposure (low = normalized difference vegetation index $\leq 0.6$ ), toxic release inventory exposure (high = residence in a county with $\geq 7$ toxic release inventory sites), heatwave day exposure (high = residence in a county with $\geq 3$ instances of heatwave days in 2018), and individual-level climate stress (high = affirmative response that climate change/global warming is stressful/makes the participant anxious).
<sup>2</sup>W1 = wave 1; W2 = wave 2. Outcome definition varied by survey wave (see Table 1).
<sup>3</sup>Adjusted for age, gender, educational attainment, and race/ethnicity [primary models].
<sup>4</sup>Adjusted for primary model covariates and discrimination experience, residence in a metropolitan region, ZIP-code level household median income, and ZIP-code level residential racial segregation.
<sup>5</sup>Stratified by race/ethnicity (non-Hispanic White or any race/ethnicity other than non-Hispanic White) and adjusted for age, gender, and educational attainment.
<sup>6</sup>Stratified by perceived discrimination experience (yes/no) and adjusted for age, gender, and educational attainment.
\*p < 0.05

In general, there was no evidence of an association between environmental context as a latent construct and any of the COVID-19 outcomes of interest (Table 6). The one exception was for associations between environmental context assessed in wave 2 with COVID-19 vaccination among participants in wave 2.

**Table 6.**
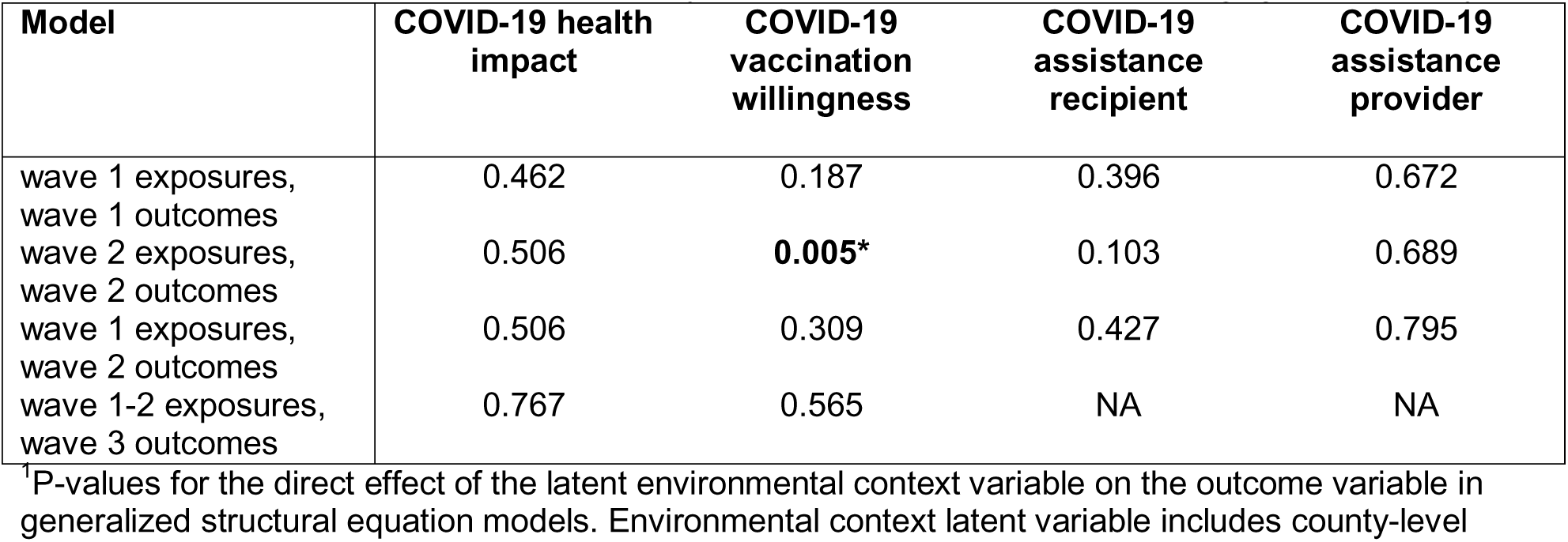
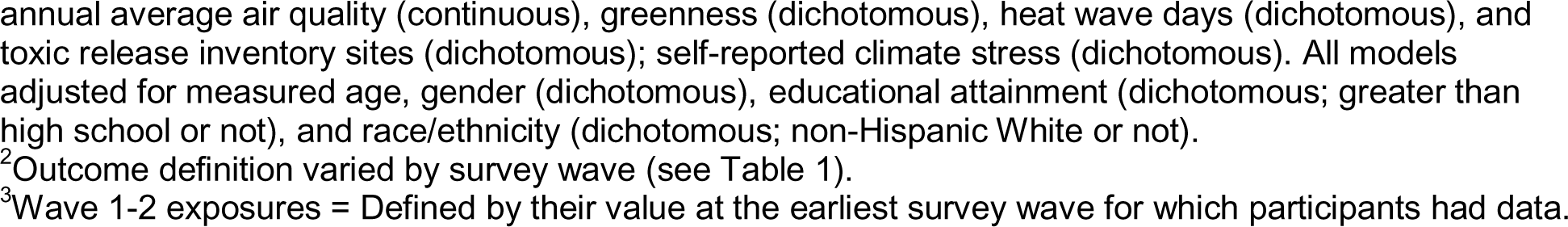
P-value for the association between environmental context^1^ and each COVID-19 outcome^2^ from the 2020-2022 Tufts Equity in Health, Wealth, and Civic Engagement Study

| Model | COVID-19 health impact | COVID-19 vaccination willingness | COVID-19 assistance recipient | COVID-19 assistance provider |
| --- | --- | --- | --- | --- |
| wave 1 exposures, wave 1 outcomes | 0.462 | 0.187 | 0.396 | 0.672 |
| wave 2 exposures, wave 2 outcomes | 0.506 | **0.005\*** | 0.103 | 0.689 |
| wave 1 exposures, wave 2 outcomes | 0.506 | 0.309 | 0.427 | 0.795 |
| wave 1-2 exposures, wave 3 outcomes | 0.767 | 0.565 | NA | NA |

<sup>1</sup>P-values for the direct effect of the latent environmental context variable on the outcome variable in generalized structural equation models. Environmental context latent variable includes county-level
annual average air quality (continuous), greenness (dichotomous), heat wave days (dichotomous), and toxic release inventory sites (dichotomous); self-reported climate stress (dichotomous). All models adjusted for measured age, gender (dichotomous), educational attainment (dichotomous; greater than high school or not), and race/ethnicity (dichotomous; non-Hispanic White or not).
<sup>2</sup>Outcome definition varied by survey wave (see Table 1).
<sup>3</sup>Wave 1-2 exposures = Defined by their value at the earliest survey wave for which participants had data.

## Discussion

Using three waves of survey data collected between 2020 and 2022 from a nationally representative sample of US adults, we found that experience of climate stress was associated with willingness to vaccinate against COVID-19 and with likelihood of receiving assistance for COVID-19. These associations were not entirely explained by political ideology or residence in a metropolitan area. We also observed that population-level environmental exposures (i.e., greenness, number of toxic release inventory sites, and number of heatwaves) were each associated with willingness to vaccinate against COVID-19. Similarly, annual PM_2.5_ exposure was associated cross-sectionally with the likelihood of provisioning assistance for COVID-19 by wave 1. Additionally, we observed that associations between certain environmental exposures and certain COVID-19 outcomes were stronger among those who identify as a race/ethnicity other than non-Hispanic White and among those who report experiencing discrimination; however, these trends were not consistent. While we doubt that certain environmental exposures (e.g., heatwave days in 2018) directly impacted COVID-19 outcomes years later, our results add to the growing body of literature suggesting that intersectional equity issues affecting likelihood of exposure to adverse environmental conditions are also associated with health-related outcomes. Our results also represent one of the first quantitative examinations into the association between environmental context and COVID-19 outcomes other than morbidity and mortality.

Our most robust observation was that people who report higher levels of climate stress were also more likely to be willing to vaccinate against COVID-19. We observed significant associations regardless of which survey wave exposures or outcomes were assessed, which covariates were included in the regression models, and whether people had experienced discrimination. This trend is not what is expected based on correlations with age group: younger people are more likely to experience climate anxiety (also termed eco-anxiety)^41^ but are less likely to be vaccinated.^42, 43^ However, the trend that people who experience more climate stress are more likely to be willing to vaccinate might be better explained by factors such as political ideology. People who vote for Democrats are more likely to vaccinate,^44, 45^ and may also be more likely to report feelings of climate stress. Notably, when we conducted a sensitivity analysis adding political ideology to the models for climate stress and vaccination willingness (Supplemental Table 1), the general trend remained the same (i.e., climate stress is positively associated with vaccination willingness, especially among people who identify as a race/ethnicity other than non-Hispanic White) but some of the associations were attenuated.

In addition to people who experience climate stress, people who were exposed to worse population-level environmental conditions (i.e., less greenness exposure, more toxic release inventory site exposure, and more heatwave day exposure) were also more likely to report a willingness to vaccinate against COVID-19. These associations for greenness exposure (but not toxic release inventory site or heatwave day exposure) were stronger among people who identify as any race/ethnicity other than non-Hispanic White. Furthermore, in wave 2, the summary environmental context variable was also significantly and positively associated with willingness to vaccinate against COVID-19. These results might be somewhat surprising since people who experience more structural inequities that likely increase adverse environmental exposures – including those who identify as part of historically marginally and oppressed communities, along with those who have lower incomes – have generally been less willing to vaccinate against COVID-19, have expressed more mistrust of COVID-19 vaccinations, and have experienced more structural barriers to vaccination.^9, 42, 46, 47^ However, it is possible that our environmental exposure variables serve as proxies for other factors, such as residence in metropolitan area (where people in metropolitan areas would be likely to have environmental exposure profiles similar to those who were more likely to be willing to vaccinate). Indeed, adjusting for residence in a metropolitan area made the associations between greenness and vaccination willingness, as well as the between heatwave days and vaccination willingness non-significant. This makes sense given that residence in an urban area is also associated with increased willingness to vaccinate against COVID-19.^48^ Additionally, it is possible that our individual environmental exposure variables and our summary environmental context variable are *not* reasonable proxies for structural factors underlying adverse environmental conditions; this could be true due to temporal variability in factors (like heatwave days) over time making 2018 estimates less useful, possible inadequacy of a preliminary statistical latent construct variable to fully capture complex interdependencies among environmental exposures, or other reasons.

In contrast to the associations, we observed between environmental exposures and vaccination willingness, we generally did not observe significant associations with COVID-19 health impacts assessed in any of the three survey waves. Although we might have expected to see significant associations based on previous studies,^22–27^ several explanations are possible for our null results. First, our definitions for COVID-19 health impact differ from many studies – especially in wave 1, where we considered possible family-member infections together with individual infections and in wave 3, where we considered delays in medical care along with individual infections. Our decision to combine delays in medical care due to COVID-19 with individual COVID-19 infections for the wave 3 definition may have conflated two different sub-populations: people who delayed medical care for themselves or their children due to COVID-19 (but not necessarily people who experienced COVID-19 infections by the third survey wave) are more likely to be better educated, have health insurance, and be at higher risk for severe COVID-19 if infected.^49, 50^ Second, selection bias might be less problematic in our study since we had a sample designed to be nationally representative and our study was not exclusively focused on COVID-19 (so people were unlikely to participate or not based on COVID-19 experiences). Third, and common to all of our analyses, there may have been exposure misclassification. This is especially true for greenness exposure, toxic release inventory site exposure, and heatwave day exposure since we dichotomized each of these population-level variables. Fourth, our measure for residential racial segregation does not accurately represent areas with low proportions of both people who identify as non-Hispanic Black and non-Hispanic White (e.g., areas with high proportions of people who identify as Hispanic or Asian) and does not capture discrimination experience. Fifth, and also common to all of our analyses, although we adjusted for a large set of covariates, it is possible we had unmeasured confounding.

As with the results for COVID-19 health impacts, we did not see consistent associations between environmental exposures and either receipt or provisioning of COVID-19 assistance. The primary exceptions were that more climate stress was associated with receipt of COVID-19 assistance by survey wave 2, and more air pollution exposure was associated with higher likelihood of provisioning COVID-19 assistance in wave 1. There was also an inconsistent trend whereby more exposure to toxic release inventory sites was associated with less likelihood of provisioning COVID-19 assistance in wave 2. Although it is a strength of our study that we were able to consider non-health COVID-19 outcomes, it is possible that there was misclassification of the outcome variables or inconsistency among participants about what they considered assistance. For example, the ways people think about a question such as *“has the government helped you deal with COVID-19 or its effects?”* might vary widely within a population or even for a given individual over time. Additionally, we understand that there is unlikely to be a direct causal relationship between the environmental exposures and these COVID-19 outcomes. To the extent that the environmental exposures serve as a proxy for the experience of environmental racism and structural oppression, we might expect to see non-causal associations. In this case, we would expect people who have been marginalized by these structural forces to be more likely to reside in areas with worse environmental exposures and perhaps to be more likely to have received COVID-19 assistance by survey wave 2 (e.g., from the federal Economic Impact Payments).^51^ We found some limited support for this hypothesis, but it was not supported by the inconsistent results observed in the analyses stratified by race/ethnicity or by discrimination experience.

Our study had several limitations. First, we previously discussed possible exposure and outcome misclassification. Additional exposure misclassification could be introduced because of our assumption that the residential location represents the appropriate spatial extent for exposure assessment, without accounting for where or when people spend time (e.g., recreation, work, study). Temporally, we assumed that our exposure measures were reasonable proxies of relative long-term environmental exposures, though we acknowledge that this assumption may be more valid for exposures like air pollution than heatwaves since there can be substantial year-to-year variability in the location and frequency of heatwave days.^52^ Similarly, and as previously discussed, some of the outcome variables (e.g., COVID-19 health impacts) represent multiple related constructs, which could plausibly have different associations with the exposures if they were not combined. This outcome misclassification could be compounded in the first wave of the survey (spring 2020) when availability and access to COVID-19 diagnostic tests were limited. Beyond exposure and outcome misclassification, our definitions for discrimination experience may also have failed to capture sufficient nuance in experience – especially since we dichotomized the variable. Similarly, our measure for residential racial segregation does not capture the full racial/ethnic diversity or discrimination experience and only focuses on racial/ethnic composition. In addition to the potential for misclassification, there were analytic limitations. For example, given the large number of comparisons we made, it is possible that some of our findings are significant due just to chance. Additionally, the latent variable for environmental context that we used in the generalized structural equation models may not fully represent participants’ environmental context, including the myriad environmental justice concerns. The results for the summary environmental context variable in particular should be considered a preliminary approach that could be further developed to understand the ways intersectional forces affect COVID-19 experiences.

Despite the limitations, our study also had several strengths. First, we used three waves of data from a nationally representative survey capturing COVID-19 experiences from 2020-2022. Second, we considered a diverse set of COVID-19 outcomes – moving beyond the well-studied associations between environmental exposures and COVID-19 incidence and mortality. Third, we considered a broad set of environmental exposures such that we could capture a more comprehensive picture of the environmental context in which participants reside. Fourth, we considered the associations between environmental context and COVID-19 from an equity lens, accounting for possible effect modification by proxies for structural inequities (i.e., race/ethnicity and discrimination experience). Finally, our environmental exposure measures were assessed for time periods prior to COVID-19 outcome occurrence.

## Conclusions

Our results add to the growing body of literature examining the intersection between environmental justice and other forms of structural oppression to suggest that environmental inequity may be associated with adverse outcomes. To our knowledge, we provide the first quantitative assessment of how certain environmental exposures (e.g., climate stress) are associated with multiple COVID-19 outcomes, including those that are not strictly health outcomes (e.g., receipt of assistance). Additionally, we provide a methodological framework that others can apply to examine the intersection between a preliminary environmental context construct, race/ethnicity (or, rather, racism), and discrimination in relation to COVID-19 or other outcomes. Finally, our results suggest that policy makers and community leaders may need to consider the role of environmental exposures in relation to pandemic preparedness and response efforts.

## Data Availability

Analytic code is available upon reasonable request from the corresponding author. Individual-level survey data may be made available upon reasonable request by email to Thomas Stopka.

## Acknowledgements

We thank all of the participants of the study.

---

**Supplemental Table 1.** Sensitivity analysis: Associations between climate stress^1^ and self-reported willingness to vaccinate from the 2020-2022 Tufts Equity in Health, Wealth, and Civic Engagement Study

|  | **Exposure: W1<sup>2</sup>**<br>**Outcome: W1**<br>OR (95% CI); p | **Exposure: W2**<br>**Outcome: W2**<br>OR (95% CI); p | **Exposure: W1**<br>**Outcome: W2**<br>OR (95% CI); p | **Exposure: W1-2<sup>3</sup>**<br>**Outcome: W3**<br>OR (95% CI); p |
| --- | --- | --- | --- | --- |
| **Climate stress** |  |  |  |  |
| Adjusted 1 <sup>4</sup> | **2.10 (1.36, 3.24);**<br>**<0.001\*** | **3.29 (2.03, 5.35);**<br>**<0.001\*** | 1.72 (0.97, 3.06);<br>0.064 | **1.79 (1.09, 2.93);**<br>**0.022\*** |
| Adjusted 2 <sup>5</sup> | 1.72 (0.91, 3.22);<br>0.092 | **3.77 (2.12, 6.73);**<br>**<0.001\*** | **3.18 (1.32, 7.67);**<br>**0.010\*** | 1.77 (0.95, 3.31);<br>0.073 |
| Non-Hispanic White<sup>6</sup> | **2.63 (1.58, 4.37);**<br>**<0.001\*** | **3.94 (1.78, 8.75);**<br>**<0.001\*** | **2.07 (1.10, 3.90);**<br>**0.024\*** | **2.32 (1.20, 4.50);**<br>**0.013\*** |
| Other than non-Hispanic White | 1.45 (0.66, 3.21);<br>0.352 | **3.35 (1.91, 5.88);**<br>**<0.001\*** | 1.25 (0.45, 3.48);<br>0.668 | 1.75 (0.82, 3.76);<br>0.149 |
| No discrimination<sup>7</sup> | **4.23 (2.07, 8.61);**<br>**<0.001\*** | **5.93 (2.62, 13.42);**<br>**<0.001\*** | **5.45 (1.77, 16.75);**<br>**0.003\*** | 1.94 (0.88, 4.26);<br>0.099 |
| Discrimination | 1.58 (0.88, 2.86);<br>0.126 | **3.09 (1.66, 5.78);**<br>**<0.001\*** | 0.96 (0.44, 2.10);<br>0.920 | **2.01 (1.08, 3.77);**<br>**0.029\*** |

<sup>1</sup>Exposure is individual-level climate stress (high = affirmative response that climate change/global warming is stressful/makes the participant anxious).
<sup>2</sup>W1 = wave 1; W2 = wave 2; W3 = wave 3. Outcome definition varied by survey wave (see Table 1).
<sup>3</sup>W1-2 = Defined by their value at the earliest survey wave for which participants had data.
<sup>4</sup>Adjusted for age, gender, educational attainment, race/ethnicity, and political affiliation [primary models].
<sup>5</sup>Adjusted for primary model covariates and discrimination experience, residence in a metropolitan region, ZIP-code level household median income, and ZIP-code level residential racial segregation.
<sup>6</sup>Stratified by race/ethnicity (non-Hispanic White or any race/ethnicity other than non-Hispanic White) and adjusted for age, gender, educational attainment, and political affiliation.
<sup>7</sup>Stratified by perceived discrimination experience (yes/no) and adjusted for age, gender, educational attainment, and political affiliation.
\*p <0.05

**Supplemental Table 2.** Associations between environmental exposures^1^ and self-reported COVID-19 health impact from the 2020-2022 Tufts Equity in Health, Wealth, and Civic Engagement Study

|  | **Exposure: W1<sup>2</sup>**<br>**Outcome: W1**<br>OR (95% CI); p | **Exposure: W2**<br>**Outcome: W2**<br>OR (95% CI); p | **Exposure: W1**<br>**Outcome: W2**<br>OR (95% CI); p | **Exposure: W1-2<sup>3</sup>**<br>**Outcome: W3**<br>OR (95% CI); p |
| --- | --- | --- | --- | --- |
| **Annual average PM<sub>2.5</sub> (µg/m<sup>3</sup>)** |  |  |  |  |
| Unadjusted | 0.99 (0.91, 1.08);<br>0.853 | 0.97 (0.87, 1.08);<br>0.570 | 1.00 (0.88, 1.14);<br>0.967 | 0.96 (0.87, 1.05);<br>0.369 |
| Adjusted 1<sup>4</sup> | 0.97 (0.89, 1.06);<br>0.481 | 0.97 (0.87, 1.08);<br>0.538 | 1.01 (0.88, 1.15);<br>0.897 | 0.98 (0.89, 1.08);<br>0.686 |
| Adjusted 2<sup>5</sup> | 0.96 (0.85, 1.08);<br>0.466 | 0.98 (0.87, 1.10);<br>0.699 | 0.99 (0.83, 1.19);<br>0.956 | 1.02 (0.91, 1.14);<br>0.712 |
| Non-Hispanic White<sup>6</sup> | 0.93 (0.83, 1.04);<br>0.202 | 0.92 (0.78, 1.08);<br>0.310 | 0.91 (0.78, 1.07);<br>0.270 | 0.99 (0.88, 1.13);<br>0.934 |
| Other than non-Hispanic White | 1.02 (0.90, 1.17);<br>0.732 | 1.01 (0.87, 1.16);<br>0.920 | 1.10 (0.87, 1.39);<br>0.435 | 0.94 (0.81, 1.08);<br>0.372 |
| No discrimination<sup>7</sup> | 0.92 (0.80, 1.05);<br>0.221 | 1.02 (0.88, 1.20);<br>0.768 | 1.13 (0.93, 1.36);<br>0.222 | 0.99 (0.86, 1.14);<br>0.924 |
| Discrimination | 1.00 (0.90, 1.11);<br>0.970 | 0.92 (0.79, 1.08);<br>0.306 | 0.93 (0.78, 1.12);<br>0.463 | 0.94 (0.83, 1.06);<br>0.324 |
| **Greenness** |  |  |  |  |
| Unadjusted | 1.24 (0.89, 1.73);<br>0.209 | 1.31 (0.85, 2.02);<br>0.222 | 1.63 (0.92, 2.90);<br>0.093 | 0.78 (0.54, 1.13);<br>0.197 |
| Adjusted 1 | 1.09 (0.76, 1.55);<br>0.648 | 1.08 (0.68, 1.73);<br>0.741 | 1.26 (0.69, 2.32);<br>0.451 | 0.80 (0.54, 1.17);<br>0.247 |
| Adjusted 2 | 1.02 (0.66, 1.58);<br>0.933 | 1.02 (0.62, 1.68);<br>0.940 | 1.30 (0.65, 2.62);<br>0.461 | 0.66 (0.42, 1.05);<br>0.080 |
| Non-Hispanic White | 1.15 (0.72, 1.84);<br>0.552 | 1.51 (0.73, 3.14);<br>0.264 | 1.53 (0.73, 3.20);<br>0.261 | 0.69 (0.40, 1.19);<br>0.181 |
| Other than non-Hispanic White | 1.02 (0.61, 1.71);<br>0.936 | 1.06 (0.62, 1.80);<br>0.829 | 1.42 (0.60, 3.38);<br>0.427 | 1.08 (0.64, 1.80);<br>0.782 |
| No discrimination | 1.61 (0.94, 2.77);<br>0.083 | 1.64 (0.89, 3.02);<br>0.111 | 2.06 (0.98, 4.36);<br>0.058 | 0.98 (0.58, 1.67);<br>0.949 |
| Discrimination | 1.05 (0.68, 1.62);<br>0.822 | 1.14 (0.61, 2.11);<br>0.681 | 1.35 (0.60, 3.08);<br>0.469 | 0.66 (0.39, 1.10);<br>0.108 |
| **Toxic release inventory sites** |  |  |  |  |
| Unadjusted | 1.29 (0.90, 1.84);<br>0.163 | 0.70 (0.43, 1.14);<br>0.147 | 0.65 (0.36, 1.20);<br>0.169 | 1.07 (0.73, 1.56);<br>0.721 |
| Adjusted 1 | 1.19 (0.83, 1.70);<br>0.344 | 0.68 (0.41, 1.13);<br>0.136 | 0.65 (0.35, 1.22);<br>0.180 | 1.08 (0.73, 1.60);<br>0.705 |
| Adjusted 2 | 1.07 (0.63, 1.82);<br>0.789 | 0.83 (0.44, 1.57);<br>0.565 | 0.94 (0.34, 2.58);<br>0.906 | 0.94 (0.56, 1.58);<br>0.824 |
| Non-Hispanic White | 1.31 (0.85, 2.00);<br>0.220 | 0.93 (0.44, 1.96);<br>0.843 | 0.91 (0.42, 1.99);<br>0.810 | 0.89 (0.53, 1.48);<br>0.647 |
| Other than non-Hispanic White | 0.98 (0.51, 1.88);<br>0.943 | **0.41 (0.22, 0.75);**<br>**0.004\*** | 0.39 (0.14, 1.10);<br>0.076 | 1.60 (0.85, 3.01);<br>0.145 |
| No discrimination | 1.29 (0.69, 2.40);<br>0.419 | 1.06 (0.50, 2.23);<br>0.875 | 1.20 (0.50, 2.91);<br>0.678 | 0.67 (0.38, 1.18);<br>0.162 |
| Discrimination | 1.20 (0.77, 1.87);<br>0.420 | **0.50 (0.26, 0.96);**<br>**0.036\*** | 0.46 (0.21, 1.02);<br>0.056 | 1.43 (0.83, 2.45);<br>0.197 |
| **Heatwave days** |  |  |  |  |
| Unadjusted | 1.20 (0.86, 1.67);<br>0.286 | 1.13 (0.71, 1.81);<br>0.601 | 1.29 (0.73, 2.28);<br>0.389 | 1.02 (0.72, 1.45);<br>0.911 |
| Adjusted 1 | 1.18 (0.84, 1.66);<br>0.334 | 1.00 (0.63, 1.59);<br>0.994 | 1.04 (0.58, 1.86);<br>0.896 | 0.96 (0.67, 1.37);<br>0.817 |
| Adjusted 2 | 1.27 (0.80, 2.01);<br>0.308 | 1.20 (0.70, 2.05);<br>0.503 | 1.45 (0.67, 3.12);<br>0.341 | 1.04 (0.68, 1.60);<br>0.864 |
| Non-Hispanic White | 1.10 (0.74, 1.62);<br>0.638 | 1.06 (0.57, 1.99);<br>0.849 | 1.11 (0.56, 2.18);<br>0.769 | 0.76 (0.47, 1.23);<br>0.269 |
| Other than non-Hispanic White | 1.21 (0.66, 2.25);<br>0.537 | 1.12 (0.60, 2.06);<br>0.727 | 1.34 (0.48, 3.70);<br>0.577 | 1.51 (0.89, 2.58);<br>0.127 |
| No discrimination | 1.04 (0.63, 1.74);<br>0.870 | 1.43 (0.73, 2.80);<br>0.291 | 1.26 (0.54, 2.95);<br>0.597 | 1.06 (0.64, 1.77);<br>0.813 |
| Discrimination | 1.28 (0.83, 1.99);<br>0.260 | 0.95 (0.51, 1.78);<br>0.883 | 1.25 (0.56, 2.79);<br>0.582 | 0.92 (0.56, 1.50);<br>0.736 |
| **Climate stress** |  |  |  |  |
| Unadjusted | 1.15 (0.86, 1.53);<br>0.337 | 0.84 (0.55, 1.30);<br>0.438 | 1.28 (0.78, 2.13);<br>0.330 | 1.10 (0.80, 1.50);<br>0.563 |
| Adjusted 1 | 1.01 (0.75, 1.36);<br>0.940 | 0.83 (0.52, 1.30);<br>0.411 | 1.26 (0.74, 2.16);<br>0.393 | 1.00 (0.73, 1.38);<br>0.979 |
| Adjusted 2 | 1.03 (0.70, 1.52);<br>0.866 | 0.75 (0.48, 1.18);<br>0.216 | 1.67 (0.80, 3.46);<br>0.171 | 0.98 (0.67, 1.44);<br>0.929 |
| Non-Hispanic White | 1.04 (0.73, 1.47);<br>0.836 | 0.95 (0.48, 1.87);<br>0.889 | 1.54 (0.84, 2.83);<br>0.159 | 0.95 (0.62, 1.46);<br>0.815 |
| Other than non-Hispanic White | 0.98 (0.58, 1.65);<br>0.938 | 0.75 (0.44, 1.28);<br>0.290 | 0.94 (0.38, 2.32);<br>0.900 | 1.04 (0.64, 1.69);<br>0.859 |
| No discrimination | 1.08 (0.67, 1.73);<br>0.763 | 0.51 (0.25, 1.01);<br>0.053 | 1.47 (0.72, 3.01);<br>0.288 | 0.78 (0.48, 1.25);<br>0.298 |
| Discrimination | 0.98 (0.67, 1.44);<br>0.932 | 1.20 (0.67, 2.16);<br>0.539 | 1.30 (0.60, 2.81);<br>0.510 | 1.11 (0.72, 1.72);<br>0.631 |

<sup>1</sup>Environmental variables include county-level annual air pollution exposure, greenness exposure (low = normalized difference vegetation index $\leq 0.6$ ), toxic release inventory exposure (high = residence in a county with $\geq 7$ toxic release inventory sites), heatwave day exposure (high = residence in a county with $\geq 3$ instances of heatwave days in 2018), and individual-level climate stress (high = affirmative response that climate change/global warming is stressful/makes the participant anxious).
<sup>2</sup>W1 = wave 1; W2 = wave 2; W3 = wave 3. Outcome definition varied by survey wave (see Table 1).
<sup>3</sup>W1-2 = Defined by their value at the earliest survey wave for which participants had data.
<sup>4</sup>Adjusted for age, gender, educational attainment, and race/ethnicity [primary models].
<sup>5</sup>Adjusted for primary model covariates and discrimination experience, residence in a metropolitan region, ZIP-code level household median income, and ZIP-code level residential racial segregation.
<sup>6</sup>Stratified by race/ethnicity (non-Hispanic White or any race/ethnicity other than non-Hispanic White) and adjusted for age, gender, and educational attainment.
<sup>7</sup>Stratified by perceived discrimination experience (yes/no) and adjusted for age, gender, and educational attainment.
\*p < 0.05

**Supplemental Table 3.**
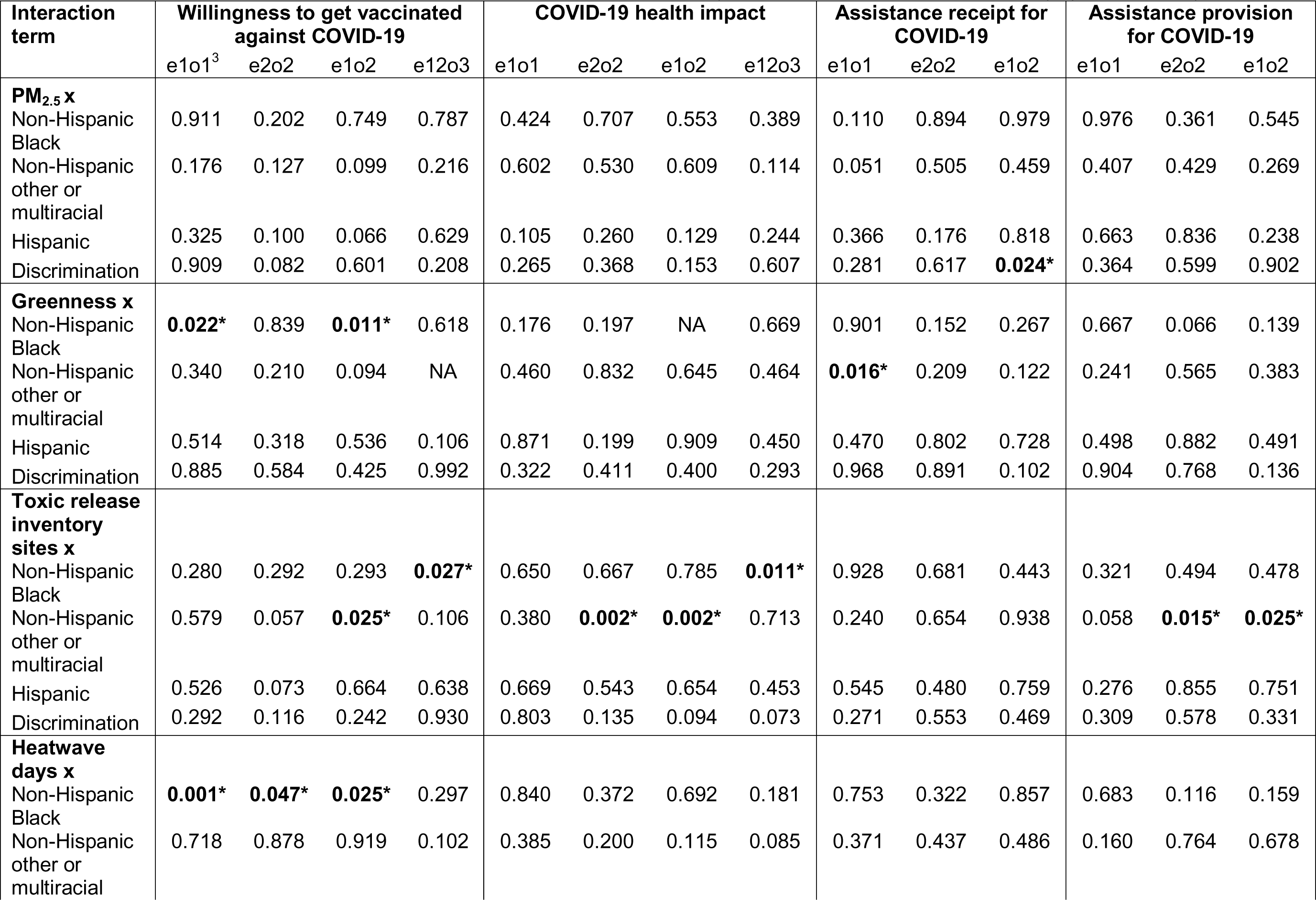
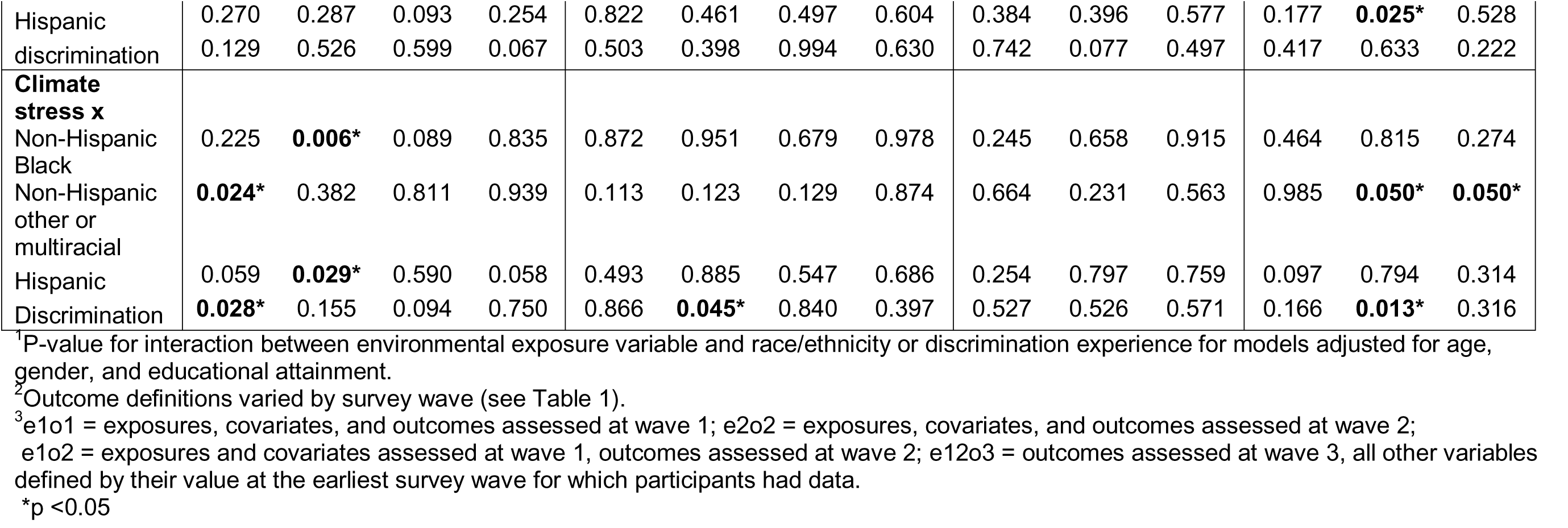
P value for interactions^1^ between environmental exposures and race/ethnicity or discrimination for their associations with COVID-19 outcomes^2^ from the 2020-2022 Tufts Equity in Health, Wealth, and Civic Engagement Study

